# A Five-Year Review of the Rates and Indications for Caesarean Section at Mbale Regional Referral Hospital: A Cross Sectional Study

**DOI:** 10.1101/2025.03.02.25323178

**Authors:** Annette Jane Mugabe Namugaya, Milton Musaba, Amongin Dinah, Gabriel Julius Wandawa, Posiano Mulalu

## Abstract

**Introduction:** Globally, there has been an increase in the caesarean section rates. Whereas, the rising caesarean section rate present a public health challenge given its implications on the individual and families amidst the limited health care resources, there is a stark paucity of data on its rate and indications in our setting. Therefore, the overarching aim of this study was to review files/charts from June 2013 to July 2018 inclusive to determine the rates and indications for caesarean section in Mbale Regional Referral Hospital hence provide evidence based recommendations to the stake holders.

**Methods:** This was a hospital cross-sectional study, based on a 5 years retrospective chart review. It was conducted in a public hospital setting where 46,575 patient files were included, of these 11,514 were caesarean section deliveries. Trained research assistants extracted data from the patient case files using a structured questionnaire. Descriptive statistics in form of frequency tables, alongside graphs, percentages, means and standard deviation were computed. Data were analyzed using STATA/SE Version 14.2.

**Results:** The rate of CS during this study was 24.7% (11,514 CS cases out of 46,575 total deliveries). Of these CS deliveries, elective CS deliveries accounted for 372/11,514 (3.2%) while 11,142/11,514 (96.8%) the emergency CS deliveries. Of the 11,514 caesarean section files, 6,466 had complete data and constituted the study sample. The commonest indications for emergency CS were Cephalo pelvic disproportion, microsomia, and fetal distress, abruptio-placenta, previous scar, and premature rupture of membranes.

**Conclusion and Recommendations:** Results show a progressively increasing CS rate from 2013 to 2018 and the common indications for CS can be screened for during Antenatal visits. This suggests that various stakeholders may need to; sensitize pregnant mothers to always attend antenatal visits, and health workers to always carefully examine pregnant mothers for such indications during antenatal checkups and advise accordingly, so as to scale down CS rate.

**Author Summary:** This study provides a detailed retrospective analysis of the Caesarean Section (CS) rates and the underlying indications at Mbale Regional Referral Hospital (MRRH) in Uganda, spanning from June 2013 to July 2018. This cross-sectional study utilizes a chart review of 46,575 patient records, with 11,514 cases of CS, to assess trends in CS rates and their associated factors. The study found a significant rise in CS rates over the five years, with an overall rate of 24.7%, exceeding the World Health Organization’s recommended threshold of 10-15%. Emergency CS deliveries were the predominant type, accounting for 96.8% of the cases, with common indications including Cephalo-pelvic disproportion, fetal distress, and previous cesarean sections.

The authors argue that while rising CS rates pose a public health challenge, the most frequent indications for CS are identifiable and manageable with early screening during antenatal visits. The study highlights the need for increased awareness and education to optimize the use of CS, suggesting that antenatal care could potentially help in reducing unnecessary CS procedures. Furthermore, it explores demographic factors, such as maternal age, parity, and educational level, and their correlations with CS rates, providing valuable insights into the socio-economic and health contexts influencing the rising CS trends in Eastern Uganda. The authors recommend targeted interventions for both healthcare providers and the community to address the root causes of high CS rates, contributing to more efficient and evidence-based obstetric care.

## Introduction

The rate at which Caesarean Section (CS) or surgical birth is being carried out in Eastern Uganda is alarming. As a result many women are delivered using this method where the rate at which CS is being performed is higher (20% -28%) than World Health Organization’s (WHO) recommended threshold of 10% - 15%. However, there is no justification for the higher caesarean section rates (Baron, 2016) and it is difficult to know an ideal CS rate for a hospital. It is not only Mbale Regional Referral Hospital (Eastern Uganda) that is experiencing a steep increase in CS rates but this is rather a global problem(Gibbons et al., 2010). There is increase is in both primary and repeat cesarean section deliveries and little research exists on “Rates and Indications for Caesarean Section” The recent statistics from 150 countries (Betran et al., 2016) “WHO Statements on Caeserean section Rates” in Brazil indicate that the average global CS rate is 18.6% of all births, meaning that 1 in 5 women around the world give birth by CS which rate is said to have increased by 12.4% (from 6.0% to 27.2%) in developed and developing regions respectively with an average annual rate of increase (AARI) of 4.4%.

At regional level, Latin America and the Caribbean had the largest absolute increases of CSR (19.4%, from 22.8% to 42.2%). However, the 42.9% CSR makes South America a sub region with the highest average CSR in the world, while Africa registered the lowest CS rates in the world with 4.5% increase (from 2.9% to 7.4%) and specifically Western Africa (3%) with the largest rise being noticed in Egypt, Tunisia and Morocco (Betrán A et al., 2016). In East Africa, countries like Tanzania, one of its referral hospital registered a rise in CSR from 29.9 % to 35.5% in the period of 2005 and 2010 (Worjoloh et al., 2012). Basing on a study conducted in Uganda, where 13 RRHs in Uganda were surveyed, it was reported that most of them had CSR above the WHO recommendation level with Mbarara RRH taking the lead (37%) followed by Hoima RRH (34%), with the least being Gulu RRH (10%). (Atuheire, 2016)The CSR where the study was conducted (MRRH) over a period of 5 years (July 2013-June 2018) indicated a significant rise from 17.3% to 31.1% (Appendices IVA – E). This CS rate was above the WHO recommended level which is 10% to 15% and therefore it formed the basis of the research.

Emergency CS was commonly performed for; fetal distress (39.3%), abruptio placenta (30.3%) and previous CS (25.6%). The maternal complication rate for women who had CS delivery during the study period was 3.2% with the main complication being PPH constituting 55.6%.The complications were noted to have been more frequent in emergency CS than in elective CS. The fetal complication rate during the study was 22.1% with the main complication being fetal asphyxia which constituted 69.8%. Parameters of fetal outcomes were determined by Apgar score at birth, birth weight and number of babies delivered. Both very low and very high CS rates are associated with adverse maternal and neonatal outcomes, high cost issues to the individual, family, community, hospital and the nation with inequitable access.(Baron, 2016).

Some few researches have been conducted; but there is scanty/no information about “Rates and indications for caesarean section in MRRH”. This study looks at the rates and indications for caesarean section in Mbale Regional Referral Hospital.

## Methods and materials

### Ethics Statement

The Ethics Review Committee of Mbale Regional Referral Hospital and Uganda National Council for Science and Technology approved the study under approval numbers “MRRH-REC OUT 055/2020” and “HS777ES” respectively. Administrative ethical clearance was obtained from Mbale Regional Referral Hospital administration where the study took place. For safety and confidentiality, when patients are discharged from the ward, their records/files were kept on the ward in the nurses’ duty rooms in cabinets or boxes which were properly labeled for some longtime like 2 years depending on the availability of space before being taken to the records office for permanent storage. Only codes were used in the Data Extraction Table (DET) not names of patients in order to secure their identity as human subjects’ rights were protected. Medical records stored on the wards did not leave the study sites and was only used for the study purpose. Analyzed data was aggregated with no individual patient identifiers. Data was only accessed by the study team. The written informed consent was waived off by the Mbale Hospital Review Ethics Committee because the study is a retrospective chart review not involving human participants.

### Study Design

A cross-sectional study involving a retrospective chart review of available records of the CS performed in MRRH (general side) from June 2013 to July 2018 inclusive. This helped to capture information at a snap shot about files and charts for mothers who delivered from MRRH (Eastern Uganda) during the study period.

### Study setting

The study was conducted in the Eastern part of Uganda in Mbale district, at MRRH (Obstetrics and Gynaecology theatre, postnatal and Maternity1 wards). MRRH is situated about 250 km Northeast of Kampala, Uganda’s capital. It is a public hospital, funded by the Uganda Ministry of Health, with a bed capacity of 400. It is a regional hub that receives patients referred from the lower health centers as well as from the periphery and provides health services from all the 16 districts with an estimated population of 2,500,000 people per year. The districts that form its catchment area include; Mbale, Manafwa, Bududa, Namisindwa, Bulambuli, Sironko, Budaka, Pallisa, Kibuku, Butebo, Butaleja, Tororo, Busia, Bukwo, Kween and Kapchorwa. However, other patients come from neighbouring districts like Bukedea and Kumi which are outside the catchment area. MRRH refers the complicated cases to the national referral hospitals (NRH) within the country (Mulago NRH and Butabika NRH). MRRH offers specialized healthcare services for the catchment area and is as well a teaching hospital for medical interns (doctors, pharmacists, anesthetists and nurses), medical students from Busitema University, Mbale College of Health Sciences students, theatre assistants and many nursing institutions.

### Content Scope

The obstetrics and gynecology department is divided into units namely; labour ward (6 delivery beds), obstetrics and gynecology theatre (1 operation bed), postnatal ward (30 beds), high dependency unit (4 beds) maternity one which is a gynecological ward (23 beds), antenatal clinic, family planning clinic and gynecological out-patient clinic. However, the study was conducted from Obs and Gynae theatre, PNW and MAT 1 at MRRH. The postnatal ward admits mothers who had spontaneous normal vaginal deliveries and caesarean section who spend 3 to 4 days on the ward while the normal deliveries are discharged after 24 hours. According to PNW records on average 15 to 20 mothers are admitted daily of which, 5 to 8 mothers undergo CS. These caesarean sections are performed from Obs and Gynae theatre. It is in MAT 1 where mothers with poor obstetric outcome like a fresh Still Birth (FSB) or Macerated Still Birth (MSB) following any type of delivery and those with other gynecological cases are admitted.

Data was reviewed from the post-natal ward and Maternity 1 at Mbale Regional Referral Hospital (general side) where all the patient’s files or records are kept for some time before being taken to the records office (Central Registry) for custody, the longitudinal maternity register (HMIS Admission Register Book) from labour suite was also looked at as it contains data for all mothers admitted for delivery in MRRH as well as the Obstetrics and Gynaecology theatre operation register book (a register for all CS performed) at MRRH

### Target population

Women of childbearing age (15 – 49 years) were the target population.

### Study Population

The study population were women of child bearing age who delivered by CS at MRRH (general side) during the study period (from July 2013 to June 2018).

### Accessible Population

All Women of child bearing age who delivered at MRRH (general side) during the study.

### Inclusion Criteria

Files and Charts of women who came to deliver at MRRH during the period of study were included in the study. The study also recorded the number of all SVDs in MRRH (general side) during the study period. Mothers from Mat 1 and PNW inclusive as their records were retrieved from the same registry and it is where mothers from MRRH (general side) who had CS deliveries are admitted.

### Exclusion criteria

Files or Charts with incomplete or missing records were excluded.

**Figure 1:**
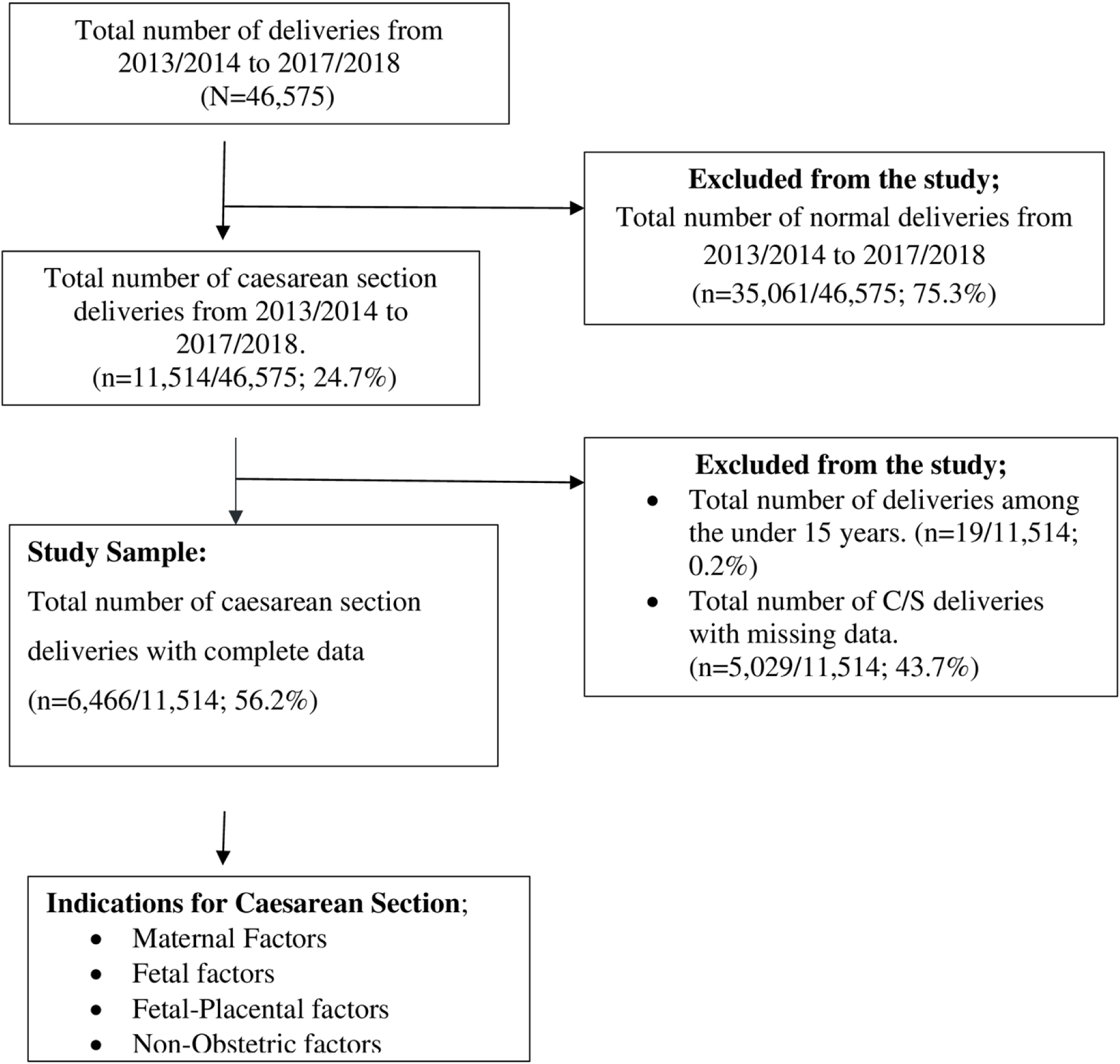
Study flow chart.

### Sample Size calculation

A total enumeration of all patients’ files was done. Those files with missing or incomplete or confusing data were eliminated from the study and those with complete data were the study sample size.

### Sampling Technique

The study consisted of reviewing Obs and Gynae theatre HMIS registers and maternal records of all mothers who delivered at MRRH during the study. This was done by reviewing the HMIS daily admission register for labour suite, looking at the daily tally sheets in labour suite, operation theatre HMIS registers from Obstetrics and Gynae theatre and admission registers from PNW.

### Study Variables

Study variables included: Dependent variables which are the number of CS performed and the outcome of the caesarean sections while independent variables are the indications for CS (maternal, fetal and obstetric factors).

### Data Collection Procedure

A pilot study was conducted prior to data collection to introduce the study to the staff responsible and to test the data collection tools; (a patient data extraction sheet for individual patients’ files, and a summary sheet of statistics of Caesarean deliveries per month for 60 months (5 years)). Training included a careful review of the variables, the data extraction form and summary sheet of statistics of CS. Any discrepancies in coding were reviewed jointly and discussed to clarify any issues.

Data were accessed for research purposes on November 20^th^, 2020 and the authors never had access to information that could identify individual participants during or after data collection.

The data collection for this study was done by the researcher as the principal investigator (PI), one trained research assistant and 3 trained midwives using a data extraction kit. All files for both CS performed and SVDs at MRRH from July 2013 to July 2018 were reviewed. This information was obtained from the Obstetrics and Gynecology operating theatre registry, labour ward records, post-natal records, maternity 1 ward records and retrieved patients’ case notes from hospital records office.

### Data Extraction Sources

Two data extraction tools were used in this study; a patient data extraction sheet for individual patients’ files, and a summary sheet of statistics of Caesarean deliveries per month for 60 months (5 years). Data was collected from the patients’ files with the variables of interest using the patient data extraction sheets which were developed by the researcher with the research objectives in mind. The patient data extraction tool had four parts. (See appendix I)

### Quality Assurance

The research assistants were trained on the questionnaire administration before commencement of the study. Prior training helped research assistants to develop competency during data collection. Every completed questionnaire was analyzed and coded by the principal investigator, checked and validated each question in each questionnaire. Logic checks were made to scrutinize for contradictory data collection.

### Data Management

Collected data was compiled, tabulated, encoded according to the variables studied by the PI and aided by the research assistant and trained health workers. In order to ensure accuracy, completeness and consistency, the research assistants had to check for completeness and accuracy before embarking on another questionnaire. Then the principal investigator had to re-check the questionnaires and ensure that there was completeness and accuracy. After which completed questionnaires were coded by the PI and data sets established with double entry checks. The collected data was entered into the computer, cleaned, processed and analyzed using STATA SE Version14.2 statistical software for analysis.

### Data Analysis

For analysis of data the variables were categorized like, maternal age (15 to 19 years, 20 to 24 years, 25 to 30 years, 31 to 34, above 35 years). The PI recorded all the deliveries (both SVDs and CSs) during the study period. The rate of CS was then be estimated by counting the number of CS performed during the study period and dividing it with the number of total deliveries within the study period, then after multiply the result by 100. The result was the percentage of CS for that study period. Descriptive analysis was made and results presented using frequency tables alongside graphs, pie charts, histograms, percentages, means and narrative texts.

After compilation of data for caesarean section at MRRH for study period, the line graph was used to show the rates of CS over the study period.

The objective two, to identify the indications for CS at MRRH, the responses on indications were sorted and coded and summarized in frequency tables, then indications with highest frequency were considered as main contributors to emerging trends in CS at MRRH.

## Results

There were 46,575 total deliveries during the 5 years period of study (July 2013 – June 2018) and in this period of study there were 11,514 cases of Caesarean Sections performed resulting into a caesarean section rate of 24.7%. Rates of CS Table 1 below shows the rates of CS during the period of study with 11,514 (24.7 %) CS cases out of 46,575 total deliveries. Of these CS deliveries, elective CS deliveries accounted for 372/11,514 (3.2%) while 11,142/11,514 (96.8%) were for the emergency CS deliveries. Of the 11,514 Caesarean section files reviewed, 6,466 files had complete data, which constituted our study sample.

**Figure 2:**
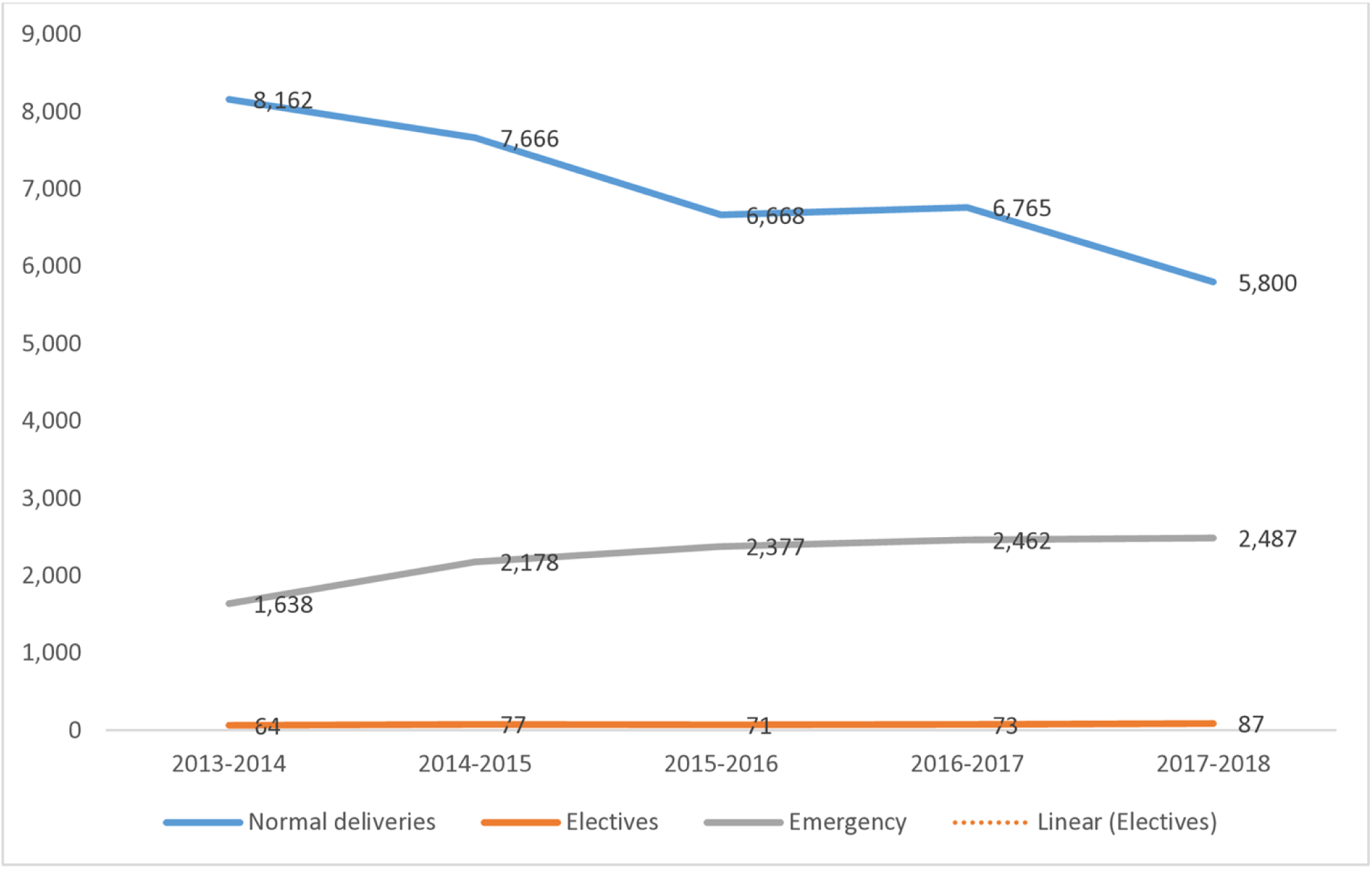
Showing the trends of all deliveries (July2013 to 2018)

**Table 1:** Summary table of mode of deliveries per year (July 2013 to June 2018)

| YEAR | NORMAL | CAESEARREANSECTION | TOTAL | C/S |
| --- | --- | --- | --- | --- |

|  | DELIVERIES | TOTAL | Electives | Emergency | DELIVERIES | rate |
| --- | --- | --- | --- | --- | --- | --- |
|  |  |  |  |  |  | (%) |
| 2013/2014 | 8,162 | 1,702 | 64 | 1,638 | 9,864 | 17.3 |
| 2014/2015 | 7,666 | 2,255 | 77 | 2,178 | 9,921 | 22.7 |
| 2015/2016 | 6,668 | 2,448 | 71 | 2,377 | 9,116 | 26.9 |
| 2016/2017 | 6,765 | 2,535 | 73 | 2,462 | 9,300 | 27.3 |
| 2017/2018 | 5,800 | 2,574 | 87 | 2,487 | 8,374 | 30.7 |
| TOTAL | 35,061 | 11,514 | 372 | 11,142 | 46,575 |  |
|  | Overall C/S rate |  |  |  |  | 24.7 |

### Demographic Factors

Out of the 11,514 files under caesarean section, 6,466 had completed data. The mean age of the 6,466 mothers was 26.74 years with a standard deviation of 5.04, and their median age was 27years (p25=23, p75=28). Others socio-demographics are shown in Table 2.

**Table 2:** Maternal Demographic characteristics.

| Demographic characteristics | Frequency, n=6,466 | Percentage (%) |
| --- | --- | --- |
| Maternal Age |  |  |
| 15 – 19 years | 555 | 8.58 |
| 20 – 24 years | 1,178 | 18.22 |
| 25 -34 years | 4,194 | 64.86 |
| 35 and above | 539 | 8.34 |
| Parity |  |  |
| Prim gravida | 3,003 | 46.44 |
| Multigravida | 2,201 | 34.04 |
| Grand multigravida | 1,262 | 19.52 |
| Race |  |  |
| White | 2 | 0.03 |
| African | 6,460 | 99.91 |
| Asian | 4 | 0.06 |
| Marital Status |  |  |
| Single | 50 | 0.77 |
| Married | 5,960 | 92.17 |
| Divorced | 19 | 0.29 |
| Separated | 166 | 2.57 |
| Widowed | 271 | 4.19 |
| Educational level |  |  |
| Unknown | 1,637 | 15.06 |
| None | 164 | 2.54 |
| Primary | 4,315 | 66.73 |
| Secondary | 1,075 | 16.63 |
| Tertiary | 912 | 14.10 |
| Employment level |  |  |
| Unemployed | 4,633 | 71.65 |
| Employed | 1,833 | 28.35 |

Of the CS performed during the study period, the majority of mothers (4,194/6,466) were between the ages of 25 to 34 years constituting 64.86%. Parity was analyzed basing on its distribution before delivery of mothers who had CS in MRRH. In this case, CS was highest among prim gravidae (first time deliveries) with 46.44% (3,003/6466) as compared to others. The majority of mothers, 6,460 (99.91%) were Africans, married were 92.17% (5,960/6,466), with a highest educational status being primary level of 66.73% (4,315/6466). The unemployed constituted 71.65% (4,633/6,466) reflecting a low socio-economic status of majority of the population that was studied as well elaborated in the histogram.

### Infant characteristics of the studied population

The characteristics of infants studied are shown in Table 3, with majority of the babies (93.88%) born by CS weighed 3 to 4 kilograms, 6,007 (90.59%) had Apgar score of 5-10, 6,310 (97.59%) babies delivered were singleton with female gender constituting 3,385 (52.35%) and first birth order 3,131 (48.42%).

**Table 3:** Birth weight of live babies who were delivered by CS during the study period.

| Birth weight(kg) | Frequency (n=6466) | Percentage |
| --- | --- | --- |
| < 2kg | 8 | 0.12 |
| 2-3 Kg | 350 | 5.41 |
| 3.1–4kg | 6,070 | 93.88 |
| >4kg | 38 | 0.59 |
| Apgar score |  |  |
| 0-5 | 624 | 9.41 |
| 5-10 | 6,007 | 90.59 |
| Number of babies delivered |  |  |
| One | 6,310 | 97.59 |
| Two | 156 | 2.41 |
| Gender of babies |  |  |
| Male | 3,081 | 47.65 |
| Female | 3,385 | 52.35 |
| Baby's birth order |  |  |
| First | 3,131 | 48.42 |
| Second | 1,150 | 17.79 |
| Third | 985 | 15.23 |
| Fourth | 550 | 8.51 |
| Fifth | 321 | 4.96 |
| Sixth | 180 | 2.78 |
| Seventh | 82 | 1.27 |
| > seventh | 67 | 1.04 |

### Indications for Cs

The registered indications for caesarean section were classified as maternal, fetal and fetal placental factors.

### Maternal Factors

The maternal indications for caesarean section recorded in this study were: previous Caesarean section, obstructed labour, Cephalic Pelvic Disproportion, APH, hypertensive disorders, multiple pregnancies, advanced maternal age, request for sterilization, Eclampsia and failed induction as shown in Table 4

**Table 4:**
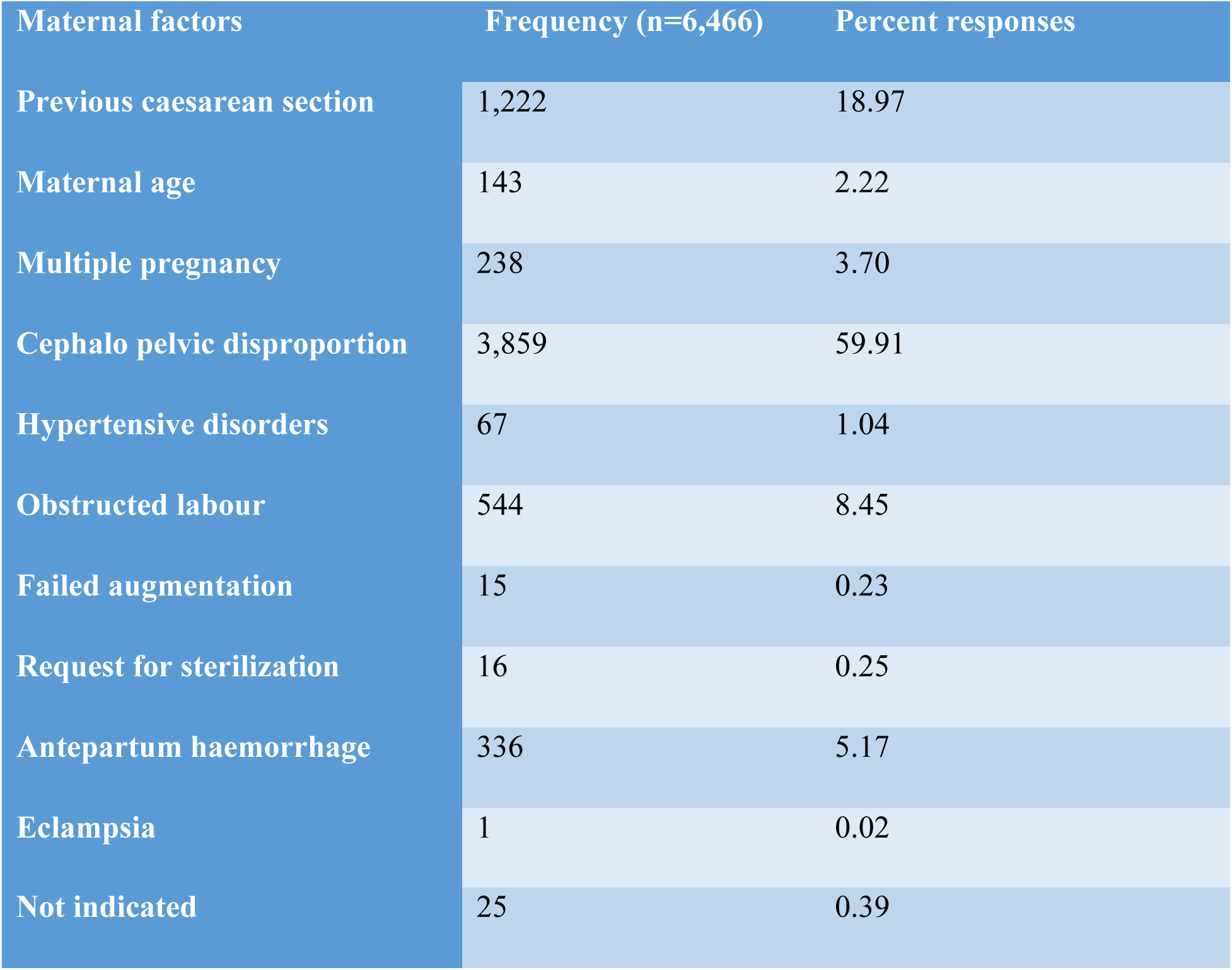
Maternal Indications for Caesarean Section.

| Maternal factors | Frequency (n=6,466) | Percent responses |
| --- | --- | --- |
| Previous caesarean section | 1,222 | 18.97 |
| Maternal age | 143 | 2.22 |
| Multiple pregnancy | 238 | 3.70 |
| Cephalo pelvic disproportion | 3,859 | 59.91 |
| Hypertensive disorders | 67 | 1.04 |
| Obstructed labour | 544 | 8.45 |
| Failed augmentation | 15 | 0.23 |
| Request for sterilization | 16 | 0.25 |
| Antepartum haemorrhage | 336 | 5.17 |
| Eclampsia | 1 | 0.02 |
| Not indicated | 25 | 0.39 |

### Fetal Factors

Table 5 shows that the major indication for caesarean section among the fetal factors were big baby (764/6466 (11.82%)) followed by fetal distress (508/6466 (7.86%)).

**Table 5:** Fetal indications for caesarean section.

| Fetal factors | Frequency (n=6,466) | Percentage of responses |
| --- | --- | --- |
| Fetal distress | 508 | 7.86 |
| Big baby | 764 | 11.82 |
| Mal-presentation | 211 | 3.26 |
| ?” | 84 | 1.30 |
| Post maturity |  |  |
| Cord prolapsed | 187 | 2.89 |
| Arm prolapsed | 59 | 0.91 |
| None | 4,653 | 71.96 |

### Fetal-Placenta Factors

Table 6 shows the fetal placental factors for CS, with premature rupture of membranes leading (69/6466 (30.3%)), followed by abruption-placenta (48/6466 (0.74%)).

**Table 6:** Fetal placental factors.

| Fetal Placental Factor | Frequency (n=6466) | Percentage responses |
| --- | --- | --- |
| Placenta previa | 11 | 0.17 |
| Abruptio-placenta | 48 | 0.74 |
| Premature rupture of membranes | 69 | 1.07 |
| Oligohydramnios | 7 | 0.11 |
| Not indicated | 6,331 | 97.91 |

### Non-Obstetric Factors

Table 7 describes non-obstetric characteristics for the Caesarean section. Majority (6,453/6466 (99.80%)) of mothers who had CS did not demand for it. Over ninety-nine percent (99.50%) did not have any medical condition associated, 6,456 /6466 (99.85%) mothers tested negative for HIV while 5,870/6,466 (90.78%) were fathers whose HIV status was negative, 6,430/6,466 (99.44%) attended ANC and 3,563/6,466 (55.10%) of mothers had no partogram use during labour. Majority 6,394/6,466 (98.89%) of the opinion for CS performed was made by intern doctors and 6,461/6,466 (99.92%) had no second opinion sought for performing caesarean section.

**Table 7:** Frequency table showing non-Obstetric factors.

| Non-obstetric factors | Frequency (n=6,466) | Percentage |
| --- | --- | --- |
| **Patient demand for caesarean section** |  |  |
| Yes | 13 | 0.20 |
| No | 6,453 | 99.80 |
| **Medical condition associated** |  |  |
| Yes | 32 | 0.50 |
| No | 6,434 | 99.50 |
| **HIV status (Mother)** |  |  |
| Positive | 10 | 0.15 |
| Negative | 6,456 | 99.85 |
| **HIV status (Father)** |  |  |
| Positive | 10 | 0.15 |
| Negative | 5,870 | 90.78 |
| Unknown | 586 | 9.06 |
| **Attended ANC** |  |  |
| Yes | 6,430 | 99.44 |
| No | 36 | 0.56 |
| Partogram use during labour |  |  |
| Yes | 2,903 | 44.90 |
| No | 3,563 | 55.10 |
| Non-obstetric factors |  |  |
|  | Frequency<br>(n=6,466) | Percentage |
| Opinion for CS made by: |  |  |
| Intern doctor | 6,394 | 98.89 |
| Medical officer | 39 | 0.60 |
| Obstetrician | 33 | 0.51 |
| Second opinion on decision for CS sought: |  |  |
| Yes | 5 | 0.08 |
| No | 6,461 | 99.92 |

### Caesarean Section Contexts

Table 8 shows that majority 6,250/6,466 (96.66%) were emergency CS, 6,270/6,466 (96.97%) had spinal anesthesia and 3,502/6,466(54.16%) had lower section type of incision made for caesarean sections.

**Table 8:** Variables that describe the context for performing Caesarean Section.

| Caesarean section | Frequency (N=6466) | Percent |
| --- | --- | --- |
| Type of operation |  |  |
| Elective | 216 | 3.34 |
| Emergency | 6,250 | 96.66 |
| Type of anaesthesia used |  |  |
| Spinal | 6,270 | 96.97 |
| General | 196 | 3.04 |
| Type of incision made |  |  |
| Abdominal | 2,964 | 45.84 |
| Lower section | 3,502 | 54.16 |

## Discussion of results

### Caesarean section rate

The caesarean section rate is an important indicator of access to essential obstetric care. This study shows an upward trend that Caesarean section rates in MRRH in 2013 - 2018 were higher than the recommended range of 10 - 15% by World Health Organization. From the previous studies as according to Atuheire et al. 2019, the CS rates in MRRH showed an increasing trend at both at facility and population levels in Uganda, stretching from 8.5% in 2012 to 11% in 2016. Our findings agree with the study by Atuheire et al. 2019, as the CS rate increased from 17% to 31% (2013 to 2018). The high prevalence rate of CS of up to 31% could be attributed to the increasing number of intern doctors, who wish also to practice Caesars, even without considering it as the last option.

There is a global concern that caesarean birth rates are increasing in the public health sector, Uganda inclusive. In the current study the mean percentage of caesarean section rate was 24.7% at MRRH. This however is not the true caesarean delivery rate as it is not reflective of the population studied because MRRH receives referrals from numerous health care facilities within and outside the catchment area. In this case, the true caesarean section rate would need to consider the number of live vaginal births occurring at Catchment area facilities.

The results in this study show that obstetric factors contributed more to the high C-section rates than non-obstetric factors in MRRH hospital. Majority, 96.7% of the C-sections were emergency procedures of these 81.0% were primary C-sections. Thus, there should be efforts in reducing primary C-section rates especially among Para 0 and Para 1 patients, and to practice to offer trial of labour to these women with previous scar who do not have any contraindications which may lead to significant reductions in MRRH C-section rates. Achieving the reductions in C-section rates in MRRH may be achieved if the obstetric guidelines and protocols are followed in assessment and management of obstetric problems

### Demographic characteristics

Caesarean sections have important long-term implications for women of reproductive age (15-49 years) yet they remain a common surgical procedure. In this study, the researcher used an approach of classifying deliveries as 15 -19 years as teen or adolescent deliveries and over 35 years as advanced maternal age. 27 years was the average age of women who underwent CS in MRRH during the period of study. The majority of caesarean sections were performed in the age group of 25-34 years, representing the most active reproductive age group. In this case, Caesarean Sections in these women may have future implications on health care by increasing the number of women with scarred uteri thereby potentially increasing future caesarean delivery rates. Ageing is linked to childbirth complication due to increased rigidity of the pelvic girdles hence higher likelihood of CS.

In this study, the majority of women who had caesarean section (66.8%) had at least attended school up to primary level and were followed with those who attained secondary education (16.7%). This implies that they had a minimum level of education (primary) and that they were not illiterate. Therefore, would ably understand and read what concerns them.

A high number of unemployed mothers were the majority (71.7%) and this may have contributed to the high rate of CS. This may imply that women who are unemployed may not be able to facilitate themselves to reach the health facilities on time as they depend on their spouses who make financial decisions. Late arrival at a health facility may complicate a pregnancy ending with caesarean section being performed. This means that spouses should be brought on board on issues concerning women especially when pregnant.

Unlike in other studies, the findings in this study is not similar in relation to parity where it was found that the increase in C-section is associated with increase in parity. In this study, the highest rate of CS was among the primigravidae with 46.4% and the least was the grand multiparous with 19.5% of the CS cases. This is probably due to the fact that women in this part of the country (Bugisu region) get pregnant at an early age in most times.

This study however described a predominantly African sample (99.9%) with the married having the highest rate (92.2%) of caesarean section performed during the study. This implies that most Africans who delivered from MRRH were married and had the capacity of getting pregnant anytime.

### Caesarean Section Context

The results show that 96.7% more than two thirds of the C-sections were emergency C-sections rather than elective procedures. Of the emergency C-sections, 81% were primary C-sections. A primary C-section has far reaching implications on subsequent deliveries. Therefore, a decision to perform a primary C-section should not be taken lightly as a local context to design applicable strategies to reduce the primary C-section rates is necessary to save a maternal, fetal and neonatal life (Belizán et al., 2018)

### Obstetric Factors

The results of this study show that the most common indications for Caesarean section was CPD, previous C-section, obstructed labour, APH, big baby, fetal distress, mal-presentation, placenta previa and abruptio placenta. The leading factor contributing to high C-section rates was CPD and this was the most common maternal indication. It was reported to be 59.9% of the C-sections performed in MRRH. This was followed by, previous scar that was recorded as another maternal indication for C-section with 19.1%. Thus, this highlights the need for medical officers to properly characterize problems in labour in Mbale regional referral hospital as an assessment of CPD implies that the woman cannot deliver vaginally with safety.

In this study, other common indications for caesarean section in particular are advanced maternal age, multiple pregnancy and hypertensive complications. The caesarean section rate at MRRH may be appropriate due to the high-risk profile of the mothers that are managed in this hospital.

### Fetal factors

Big baby was diagnosed in 764 patients (11.8% of patients) in this study. This was the leading fetal factor among the indications for emergency CS. However, universally, fetal distress is the commonest indication for emergency C-section. In this case, a big baby will depend on the size of the pelvis and outlet of the mother as CPD was the commonest maternal indication for CS in MRRH. In other words, what constitutes big baby may differ from one clinician to another basing on inter and intra-observer differences in the interpretation. Bigger babies make the delivery process to get more difficult as the size of the mother’s bony pelvis remains constant. In this case, mothers should be counseled and encouraged to attend ANC so that the weight of the babies is monitored in utero by a skilled health worker and advised accordingly before labour starts.

### Non-Obstetric Factors

Researchers in other countries attributed a sizeable proportion of Caesarean births to non-obstetric factors but in this study, non-obstetric factors did not play a major role in the high C-section rates.

This study showed that maternal request for C-section was not a major factor in increasing C-section rates as it constituted 0.2% of the C-section performed during the period of study. Indeed, it was only 13 out of 6,466 patients with CS performed that were registered as having demanded for caesarean section to be performed during the study period.

The heavy work load may make it such that even if a patient would desire a C-section with no medical indication, the patient’s desire may not be denied and not be documented too.

Also, the heavy work load makes it such that even if a patient would desire a C-section, if there was no medical indication, the patient’s desire may not be documented and would be denied.

In this study partogram was used in 44.9% of the patients who had Caesarean sections. Guidelines for management of labour recommend use of partogram to record and monitor the progress of labour. C-section rates may decrease in MRRH if partogram use is improved. However, failure to use a partogram may have been attributed to staff shortage and work overload as this is a regional referral hospital receiving complicated maternity cases.

The HIV positive mothers constituted to 0.25% of the rate of Caesarean sections performed during the study period while the negative constituted 99.8%. In this case, most HIV infected pregnant women are delivered by C-section because of the protective effect of caesarean section on mother to child transmission of HIV or if there is any other obstetric factor like previous C-section. However, it was not recorded whether women were offered elective or emergency caesarean sections basing on their HIV status.

The researcher noted that, assessment of patients and making decisions relied more on intern doctors as they constituted 98.9% of the decisions, they made to perform C-sections followed by the medical officers with 0.6%. This may explain why there were more emergency C-sections in MRRH as intern doctors try to perfect their skills in performing C-sections. According to the records, seeking of second opinion before deciding to perform a C-section (a practice known to reduce C-section rates) was not practiced in MRRH. This may explain why the increasing rate of caesarean section in MRRH.

### Strengths of the Study

The strength of this study was that data was readily available and was extracted from a database by trained health staff in a real clinical setting where the research assistants work within the department and this was after seeking permission from the ward in-charge. Sources of data were reliable as they were hospital patient records not population surveys. This eliminated errors due to participants recall and errors due to coding of deliveries as emergency or elective C -sections. Other strength of the study is that variables were described in relation to the context of caesarean section. However, this method employed reduces the possibility of sampling bias and thus enabling statistical deductions to be made on data analysis.

### Limitations of the study

Researches based on secondary data suffer from incompleteness and unreliable information. Thus, incomplete recording of all subjective issues in patients’ files was the main limitation of the study. Another limitation was the inability to fully investigate the effects of non-obstetric factors on C-section rates as the necessary information was not usually recorded in patient files. The fact that it involved one hospital was a weakness. However, there is no reason to believe that the situation in most labour suites will be different from the situation described and thus the possibility of bias affecting the results of this study must be noted. Characteristics differ in women who had trial of labour (ToL) by choice at the hospital or whose labour started at home or were referred from other health facilities after ToL from those women who underwent emergency C-section, and these differences might affect maternal and perinatal outcomes. Another challenge was in identifying complications caused by the procedure itself from those caused by the condition that required caesarean delivery. The health status of the mothers in this study may have been a determinant for caesarean delivery as well as a determinant of health outcomes. Referral cases might overestimate the true magnitude of Cesarean Section, as this is a referral hospital.

## Future Research

Future research should be considered into alternative methods of fetal monitoring mainly using a partogram for high-risk patients and its effects on caesarean section rate as well as quality improvement in data management.

## Conclusion

The rate of CS during this study was 24.7% (11,514 CS cases out of 46,575 total deliveries). Of these CS deliveries, elective CS deliveries accounted for 372/11,514 (3.2%) while 11,142/11,514 (96.8%) the emergency CS deliveries. Of the 11,514 caesarean section files, 6,466 had complete data and constituted the study sample.

The commonest indications for emergency CS were Cephalo pelvic disproportion, microsomia, and fetal distress, abruptio-placenta, previous scar, and premature rupture of membranes.

## Recommendations

Results show a progressively increasing CS rate from 2013 to 2018 and the common indications for CS can be dealt with during Antenatal visits. This suggests that various stakeholders may need to; sensitize pregnant mothers to always attend antenatal visits, health workers to always carefully examine pregnant mothers for such indications during antenatal checkups, and Health workers to advise pregnant mothers accordingly, so as to scale down CS rate.

## Data Availability

All relevant data are within the manuscript and its Supporting Information files.

## Notes

### Competing Interest Statement

The authors have declared no competing interest.

### Funding Statement

The author(s) received no specific funding for this work.

### Author Declarations

Ethics Review Committee of Mbale Regional Referral Hospital and Uganda National Council for Science and Technology

## References

1. Atuheire. (2016). Trends of Caesarean Sections in Uganda : 2012-2015, 2012–2015.

2. Baron, M. Y. (2016). Does the 10-15% Caesarean Section Rate Threshold endorsed by the World Health Organization in 1985 still apply to Modern Obstetrics in Developed Countries? The “Ideal” Caesarean Section Rate and the Stillbirth and Neonatal Death Perspective. iMedPub Journals, 2 (1), 13.

3. Belizán, Mincka Ns, Elizabeth M McClure, Sarah Saleem, Janet L Moore, Goudar S, … Patricia L Hibberd, P. M. B., Robert L Goldenberg. (2018). An approach to identify a minimum and rational proportion of caesarean sections in resource-poor settings: a global network study.

4. Betrán A, P., Ye J, Moller A-B, Zhang J, Gülmezoglu AM, & MR, T. (2016). The Increasing Trend in Caesarean Section Rates: Global, Regional and National Estimates: 1990-2014. PLoS ONE, 11(2).

5. Betran, A., P,, Torloni, M., Zhang, J., Gülmezoglu, A., Section, W. W. G. o. C., & Aleem, H., … Carroli, G. . (2016). WHO Statement on caesarean section rates. . BJOG: An International Journal of Obstetrics & Gynaecology, 123(5), 667-670.

6. Gibbons, Luz, Belizán, José M, Lauer, Jeremy A, … Fernando. (2010). The global numbers and costs of additionally needed and unnecessary caesarean sections performed per year: overuse as a barrier to universal coverage.

7. Worjoloh, A., Manongi, R., Oneko, O., Hoyo, C., Daltveit, A. K., & Westreich, D. (2012). Trends in cesarean section rates at a large East African referral hospital from 2005-2010. Open Journal of Obstetrics and Gynecology, 2(03), 255.

